# Multicenter cohort study of children hospitalized with SARS-CoV-2 infection

**DOI:** 10.1101/2021.02.19.21251340

**Authors:** Michelle Barton, Jesse Papenburg, Rolando Ulloa-Gutierrez, Helena Brenes-Chacon, Adriana Yock-Corrales, Gabriela Ivankovich-Escoto, Alejandra Soriano-Fallas, Marcela Hernandez-de Mezerville, Ari Bitnun, Shaun K. Morris, Tala El Tal, E. Ann Yeh, Peter Gill, Ronald M. Laxer, Alireza Nateghian, Behzad Haghighi Aski, Ali Manafif, Marie-Astrid Lefebvre, Chelsea Caya, Suzette Cooke, Tammie Dewan, Lea Restivo, Isabelle Viel-Thériault, Adriana Trajtman, Rachel Dwilow, Jared Bullard, Manish Sadarangani, Ashley Roberts, Nicole Le Saux, Jennifer Bowes, Jacqueline K. Wong, Rupeena Purewal, Janell Lautermilch, Kirk Leifso, Cheryl Foo, Leigh Anne Newhook, Ann Bayliss, Dara Petel, Joan Robinson, on behalf of the Paediatric Investigators Collaborative Network on Infections in Canada (PICNIC)

**Affiliations:** Department of Pediatrics, Western University, London, Ontario; Department of Pediatrics, McGill University, Montreal, Quebec; Department of Pediatrics, Hospital Nacional de Niños “Dr. Carlos Sáenz Herrera”, Caja Costarricense de Seguro Social (CCSS); San José, Costa Rica; Department of Pediatrics, University of Toronto, Toronto, Ontario; Department of Pediatrics, Iran University of Medical Sciences, Tehran, Iran; Department of Pediatrics, University of Calgary, Calgary, Alberta; Department of Pediatrics, Laval University, Quebec City, Quebec; Department of Pediatrics, University of Manitoba, Winnipeg, Manitoba; Vaccine Evaluation Center, BC Children’s Hospital Research Institute, Vancouver, BC; Department of Pediatrics, University of British Columbia, Vancouver, BC; Department of Pediatrics, University of Ottawa, Ottawa, Ontario; Department of Pediatrics, McMaster University, Hamilton, Ontario, Canada; Department of Pediatrics, University of Saskatchewan, Saskatoon, Saskatchewan; Department of Pediatrics, Queen’s University, Kingston, Ontario; Department of Pediatrics, Memorial University, St John’s, Newfoundland and Labrador; Department of Pediatrics, Trillium Health Partners, Mississauga, Ontario; Department of Pediatrics, University of Alberta, Edmonton, Alberta

**Keywords:** COVID-19, SARS-CoV-2, multi-system inflammatory syndrome in children, pediatric, disease severity, hospitalization

## Abstract

**Background:** A cohort study was conducted to describe and compare the characteristics of SARS-CoV-2 infection in hospitalized children in three countries.

**Methods:** This was a retrospective cohort of consecutive children admitted to 15 hospitals (13 in Canada and one each in Iran and Costa Rica) up to November 16, 2020. Cases were included if they had SARS-CoV-2 infection or multi-system inflammatory syndrome in children (MIS-C) with molecular detection of SARS-CoV-2 or positive SARS-CoV-2 serology.

**Results:** Of 211 included cases (Canada N=95; Costa Rica N=84; Iran N=32), 103 (49%) had a presumptive diagnosis of COVID-19 or MIS-C at admission while 108 (51%) were admitted with other diagnoses. Twenty-one (10%) of 211 met criteria for MIS-C. Eighty-seven (41%) had comorbidities. Children admitted in Canada were older than those admitted to non-Canadian sites (median 4.1 versus 2.2 years; p<0.001) and less likely to require mechanical ventilation (3/95 [3%] versus 15/116 [13%]; p<0.05). Sixty-four of 211 (30%) required supplemental oxygen or intensive care unit (ICU) admission and 4 (1.9%) died. Age < 30 days, admission outside Canada, presence of at least one comorbidity and chest imaging compatible with COVID-19 predicted severe or critical COVID-19 (defined as death or need for supplemental oxygen or ICU admission).

**Conclusions:** Approximately half of hospitalized children with confirmed SARS-CoV-2 infection or MIS-C were admitted with other suspected diagnoses. Disease severity was higher at non-Canadian sites. Neonates, children with comorbidities and those with chest radiographs compatible with COVID-19 were at increased risk for severe or critical COVID-19.

**Main points:** Approximately half of hospitalized children with laboratory confirmed MIS-C or SARS-CoV-2 infection were admitted with another primary diagnoses. The severity of disease was higher in the middle income countries (Costa Rica and Iran) than in Canada.

## Background

The coronavirus disease 2019 (COVID-19) pandemic caused by the Severe Acute Respiratory Syndrome coronavirus-2 (SARS-CoV-2) was designated by the World Health Organization (WHO) as a pandemic on March 11, 2020. It has spread worldwide with an unprecedented impact on the global health care system, with over 106 million confirmed cases and 2.3 million deaths as of February 9, 2021 [1].

Previous publications consistently described less severe clinical manifestations of COVID-19 in children as compared to adults [2, 3]. It has been postulated that protective factors in children may include a more robust innate immune response, partial immunity from prior infection with seasonal coronaviruses, and/or the lower incidence of comorbidities in children [3]. Children younger than one year of age appear to be at increased risk of hospitalization compared to older children [3, 4]. It is not clear whether this is because they have more severe disease or because the threshold for admission is lower for infants. Comorbidities increase the risk of pediatric hospitalization [5] and critical disease [6] but socio-demographic risk factors for hospitalization or critical disease in children are yet to be described.

It is estimated that at least one-third of adults with SARS-CoV-2 infection remain asymptomatic [7]. An even higher percentage of children may be asymptomatic [8]. Although neither symptomatic nor asymptomatic hospitalized children have been reported to be a source of hospital-acquired SARS-CoV-2 infections, it is possible that if proper precautions are not instituted prior to confirmation of infection, other patients, caregivers or health care workers could be exposed. Furthermore, infected caregivers accompanying their hospitalized children may serve as vectors of transmission in paediatric healthcare settings.

At the more severe end of the clinical spectrum, a small percentage of children develop a multi-system inflammatory syndrome (MIS-C) approximately 2 to 6 weeks following SARS-CoV-2 infection [9]. A systematic review of 655 MIS-C cases reported that 447 required intensive care (68%) and 11 died (1.7%) [10].

The objectives of this study were to describe and compare the characteristics and outcomes of a cohort of hospitalized children with SARS-CoV-2 infection or MIS-C in three countries, one of which is high income (Canada) and two of which are middle income (Costa Rica and Iran) [11] and to describe risk factors for severe or critical COVID-19.

## Methods

Fifteen pediatric hospitals (13 in Canada and one each in Tehran, Iran and San José, Costa Rica) entered consecutive symptomatic and asymptomatic children up to 17 years of age admitted February 1, 2020 through November 16, 2020 with laboratory-confirmed SARS-CoV-2 infection (defined by detection of SARS-CoV-2 by molecular testing from any site and/or positive serology for SARS-CoV-2), including those with MIS-C as defined by WHO criteria [9]. Those who met WHO MIS-C criteria but did not have laboratory-confirmed SARS-CoV-2 infection were excluded to ensure that only cases due to SARS-CoV-2 were included. There were no other exclusion criteria. Indications for testing and assays used varied over time and by hospital. Cases were identified through the microbiology laboratory, infection prevention and control and/or COVID-19 unit databases.

Study data were collected and managed using REDCap electronic data capture tools hosted at the University of Alberta. Ethics approval was obtained from the following ethics review boards: Comité Ético Científico Hospital Nacional de Niños, San José, Costa Rica (CEC-HNN-030-2020), Iran University of Medical Sciences Ethics Review Committee (IR.IUMS.REC.1399.187), The Hospital for Sick Children Research Ethics Board (#1000070091), Pediatric Panel of the Research Ethics Board of the Research Institute of the McGill University Health Centre (#MP-37-2021-6561), Conjoint Health Research Ethics Board, University of Calgary (REB20-0594), Children’s Hospital of Eastern Ontario Research Ethics Board (CHEOREB# 20/32X), University of British Columbia Children’s and Women’s Research Ethics Board (# H20-00977), Health Research Ethics Board, University of Manitoba (HSC23858), University of Saskatchewan Biomedical Research Ethics Board (Study 1921), Hamilton Integrated Research Ethics Board, Centre Hospitalier Universitaire de Québec-Université Laval (37-2021-6561), and Health Research Ethics Board, University of Alberta (Pro00099426). Data were collected by chart review on primary reason for admission, demographics, comorbidities, clinical presentation and course, coinfections, treatments and complications.

The STrengthening the Reporting of OBservational studies in Epidemiology (STROBE) guidelines were followed [12].

### Bias

All eligible children were included at all sites to avoid selection bias.

### Definitions

Children who were symptomatic but did not require supplemental oxygen were classified as having mild disease. Severe disease was defined as the need for supplemental oxygen due to COVID-19 or MIS-C without either death or admission to ICU. Critical disease was defined as death or admission to an ICU (excluding children admitted to ICU for other indications). The Canadian Nosocomial Infection Surveillance Program definition was used to define healthcare-associated infection: Symptom onset equal to or greater than 7 calendar days after admission to hospital, using best clinical judgement if onset is sooner [13]. Viral co-infection was defined as laboratory detection of any virus concurrent with SARS-CoV-2 infection. Bacterial co-infection was defined as laboratory detection of one or more bacteria that was treated with antibiotics by the attending physician. Testing for coinfection was at the discretion of the attending physician. Medical complexity was defined as presence of a tracheostomy, feeding tube, home supplemental oxygen, or invasive or non-invasive ventilation prior to admission. Neurological complications were defined as worsening or new onset of seizure, encephalopathy, dystonia, chorea, athetosis, hemiparesis and/or abnormal cerebrospinal fluid cell count for age. Hematologic complications were defined as disseminated intravascular coagulation, thrombosis and/or bleeding.

### Data analysis

Data were analyzed using Epi-Info™ Version 7.2. Baseline and demographic characteristics were summarized using standard descriptive statistics. In the descriptive analysis two and three-way comparisons of demographic, clinical, laboratory and treatment data as well as outcomes were done to identify differences between children from the three participating countries. Categorical data were compared using chi square or Fisher exact test and continuous data compared using ANOVA or t tests for means and Kruskall Wallis for medians. Bonferroni adjustments were made for multiple comparisons. Data on demographics and outcomes were compared for Canadian versus other sites (Iran and Costa Rica).

Logistic regression analysis was performed using R version 3.5.2 (R Core Team, Vienna, Austria) to explore predictors of severe or critical disease, comparing those with severe or critical disease to those with asymptomatic or mild disease. Adjusted odds ratios (aOR) and associated 95% confidence intervals (95% CI) were calculated. Children with MIS-C were excluded from this analysis since the risk factors for severe or critical MIS-C likely differ from those for severe or critical acute COVID-19. Children in ICU for indications judged to be unrelated to COVID-19 were also excluded. Baseline characteristics of interest included biological sex, age, country, timing of admission (March-June versus July-November [14]), and presence of one or multiple comorbidities (as listed in Table 1). Obesity and pulmonary disease (including asthma) were specifically analyzed as they are common in childhood. Clinical variables that were explored included presence of a coinfection, shortness of breath, fever for more than 5 days, presence of gastrointestinal symptoms (defined as vomiting, diarrhea or abdominal pain), chest imaging compatible with COVID-19 (the physician completing the case report form determined whether the report of the chest imaging on the chart fit with COVID-19), or abnormal laboratory markers including leukocytosis (>15 × 10^9^/L), leukopenia (<4 × 10^9^/L), neutropenia (<1.5 × 10^9^/L), lymphopenia (< 1.0 × 10^9^/L), thrombocytopenia (< 100 × 10^9^/L), thrombocytosis (> 450 × 10^9^/L), C-reactive protein above 50 mg/L, ferritin above 500 mcg/L or albumin below 29 g/L. Univariable analysis was conducted and variables with p-value ≤ 0.2 were added to the multivariable logistic regression model in a stepwise manner.

**Table 1.**
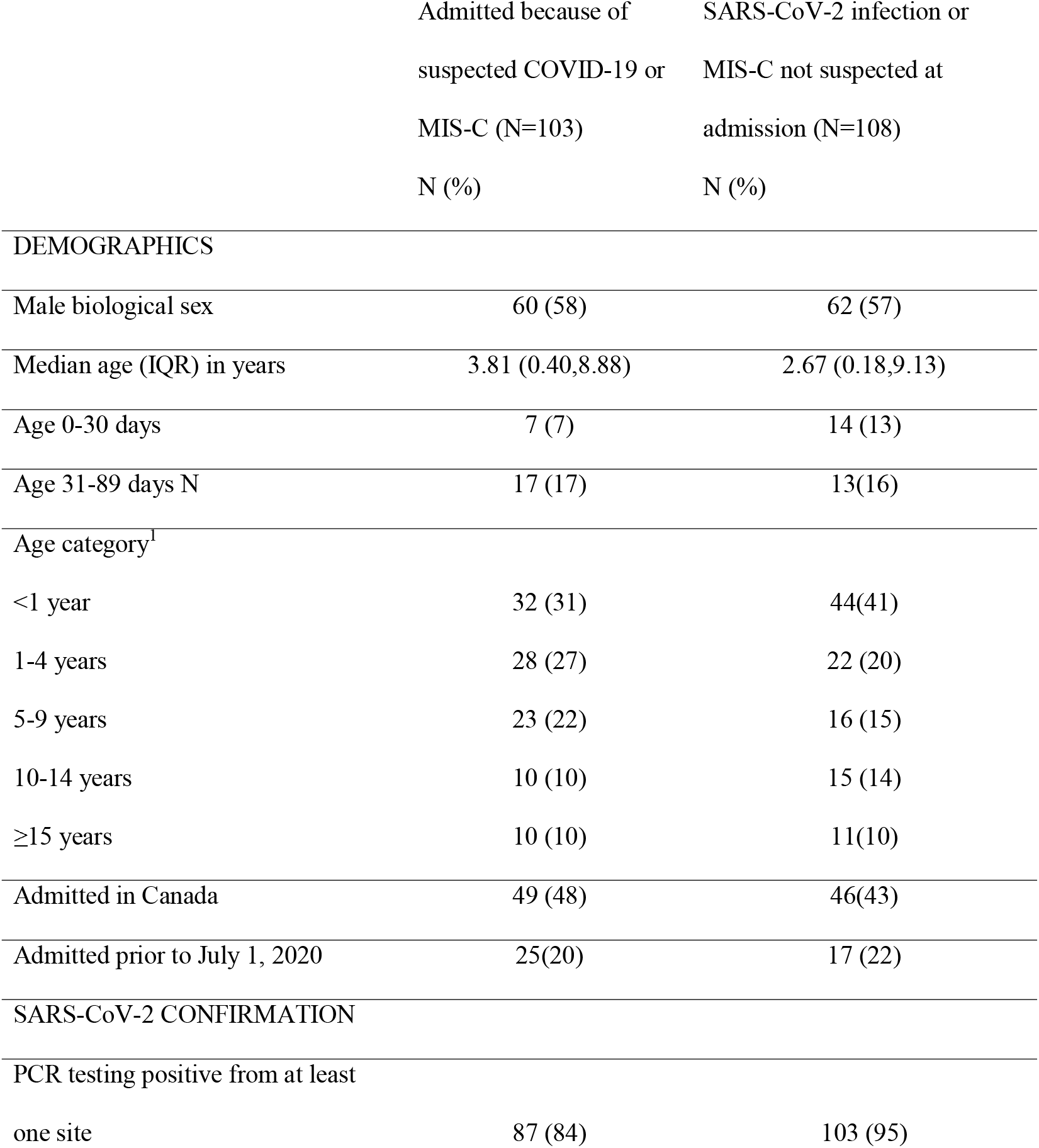
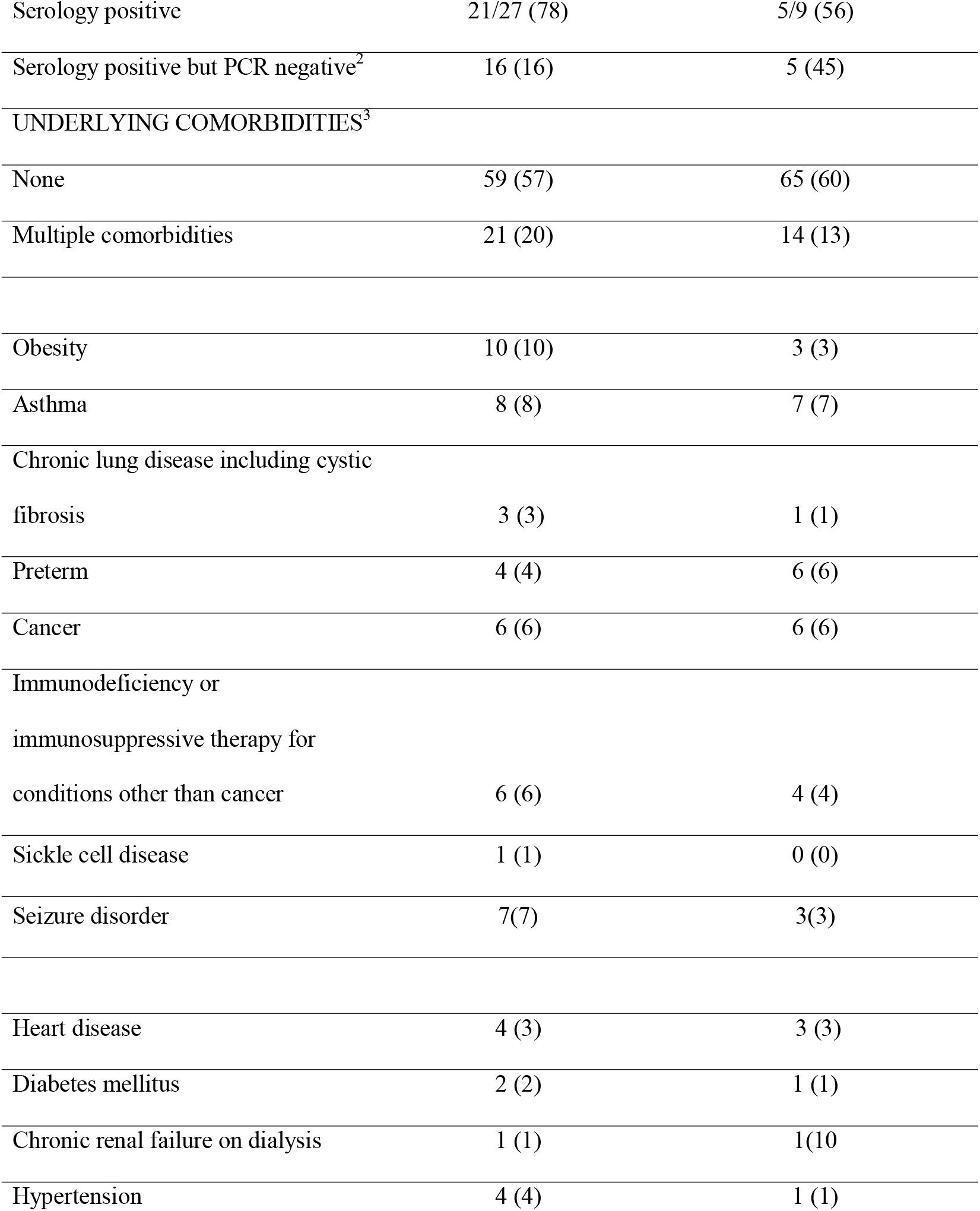
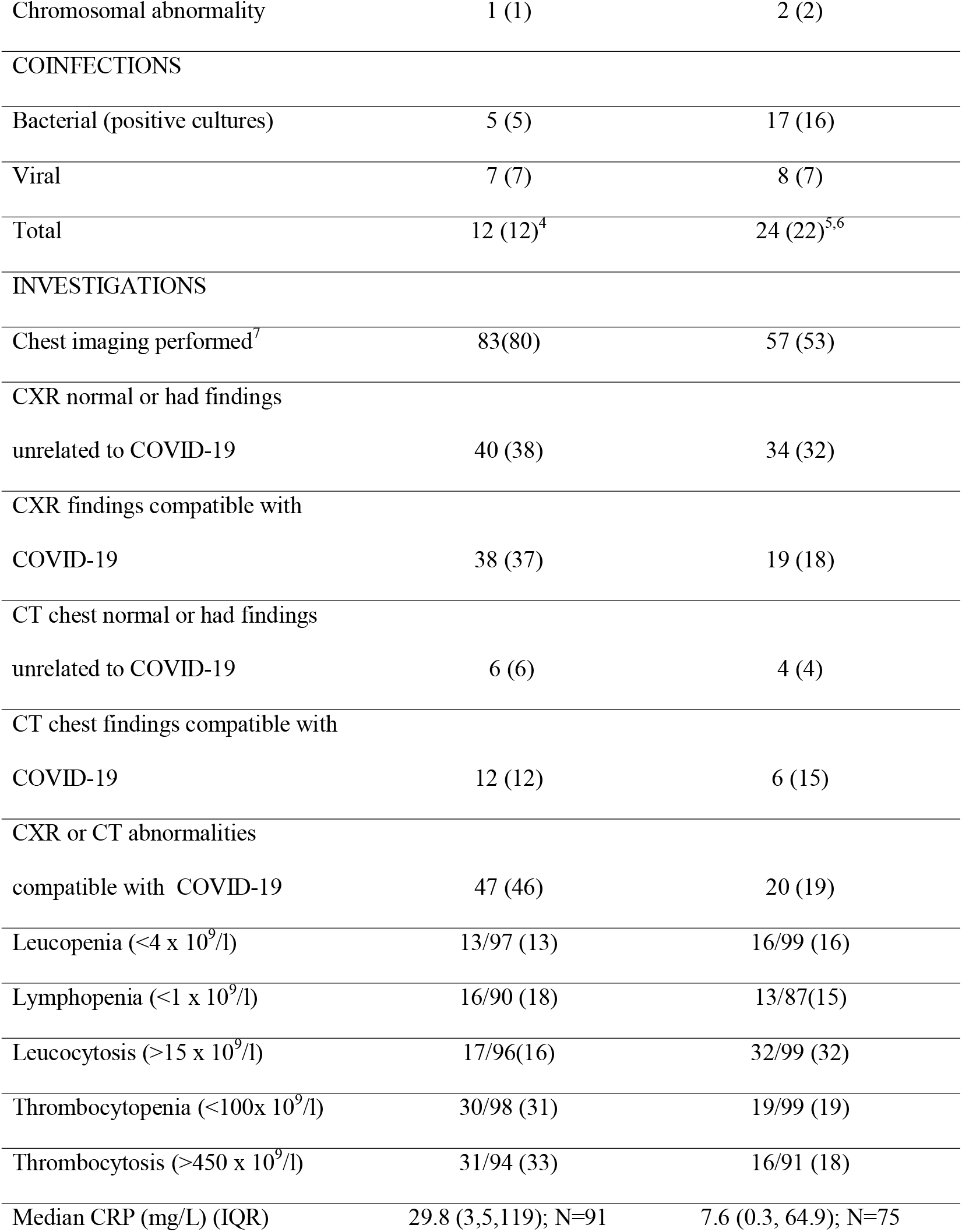
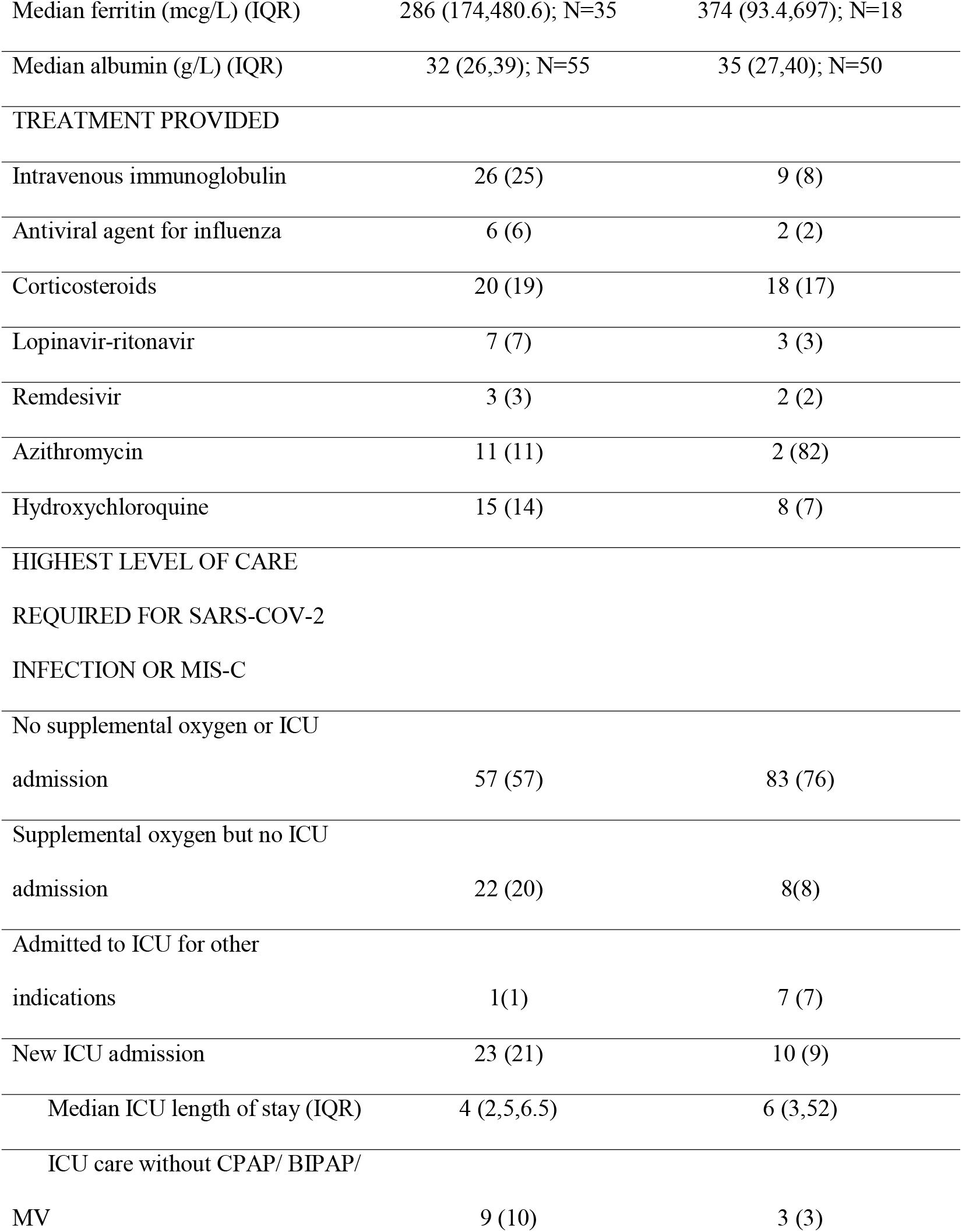
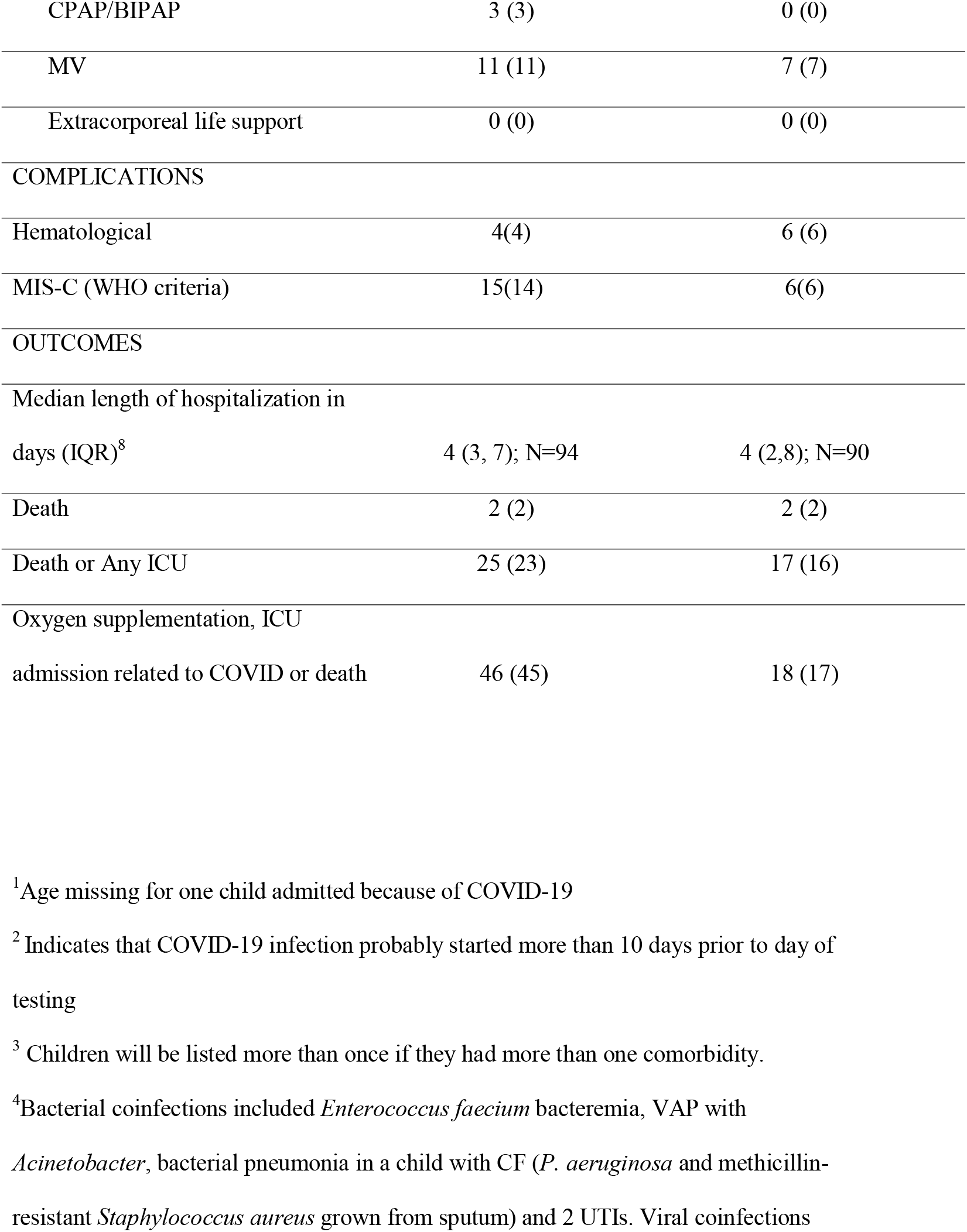
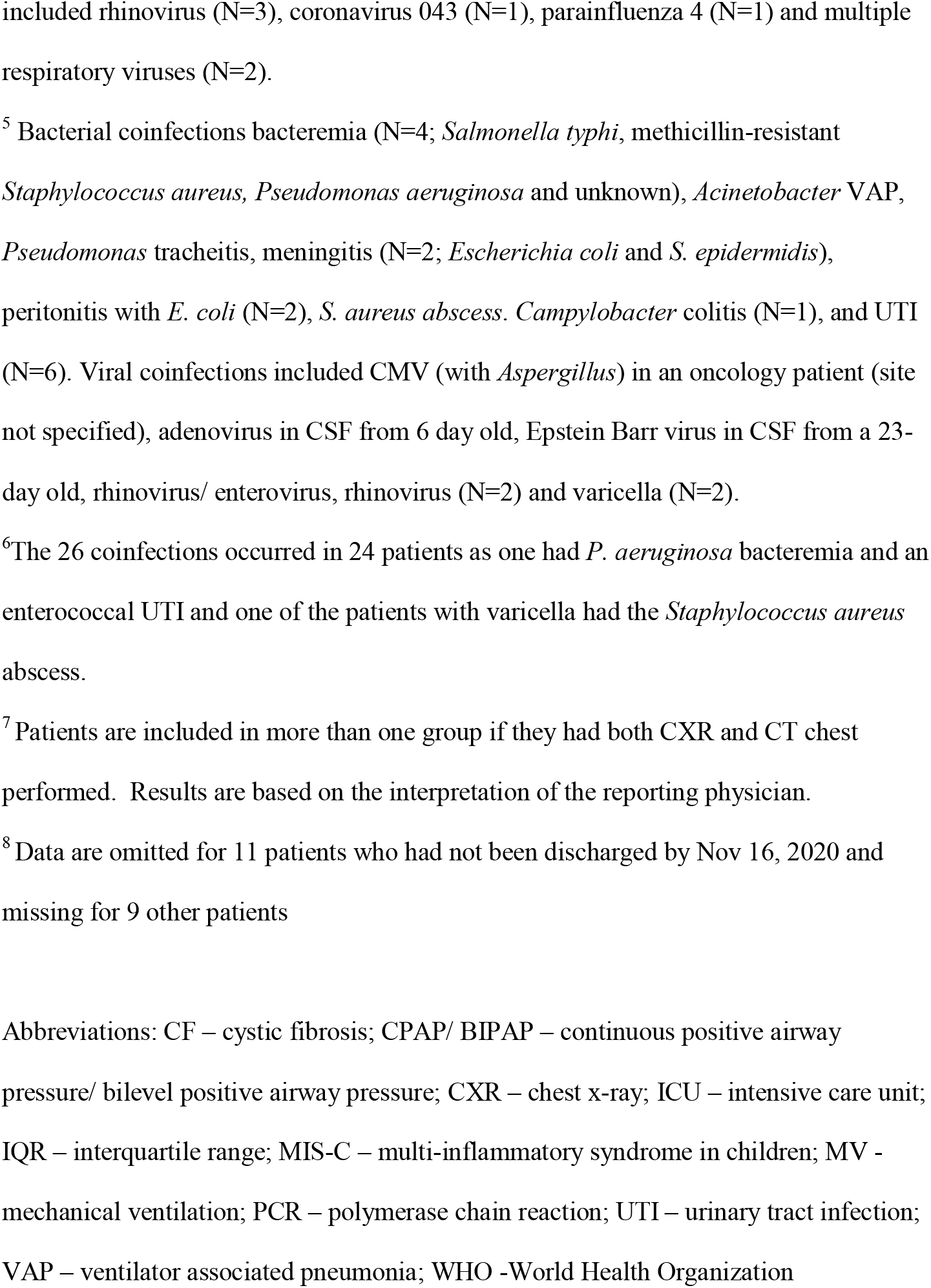
Demographic and clinical features of 211 children with SARS-CoV-2 infection or MIS-C during hospitalization

## Results

Three centers with a low community prevalence of SARS-CoV-2 infection had no patients that met the inclusion criteria. Of 294 cases enrolled from the other 12 centres, 21 with MIS-C were included while another 83 were excluded as they did not have confirmed SARS-CoV-2 infection (Figure 1). Eight (4%) of the 211 cases met the definition for healthcare-associated SARS-CoV-2. Symptom onset ranged from day 7 to day 47 (N=7; one remained asymptomatic) with viral detection ranging from day 8 to day 55. One of the 8 was an infant with extreme prematurity born to a mother with COVID-19 at delivery; the infant had previously tested negative but tested positive on day 18 of life when new respiratory symptoms developed.

**Figure 1.**
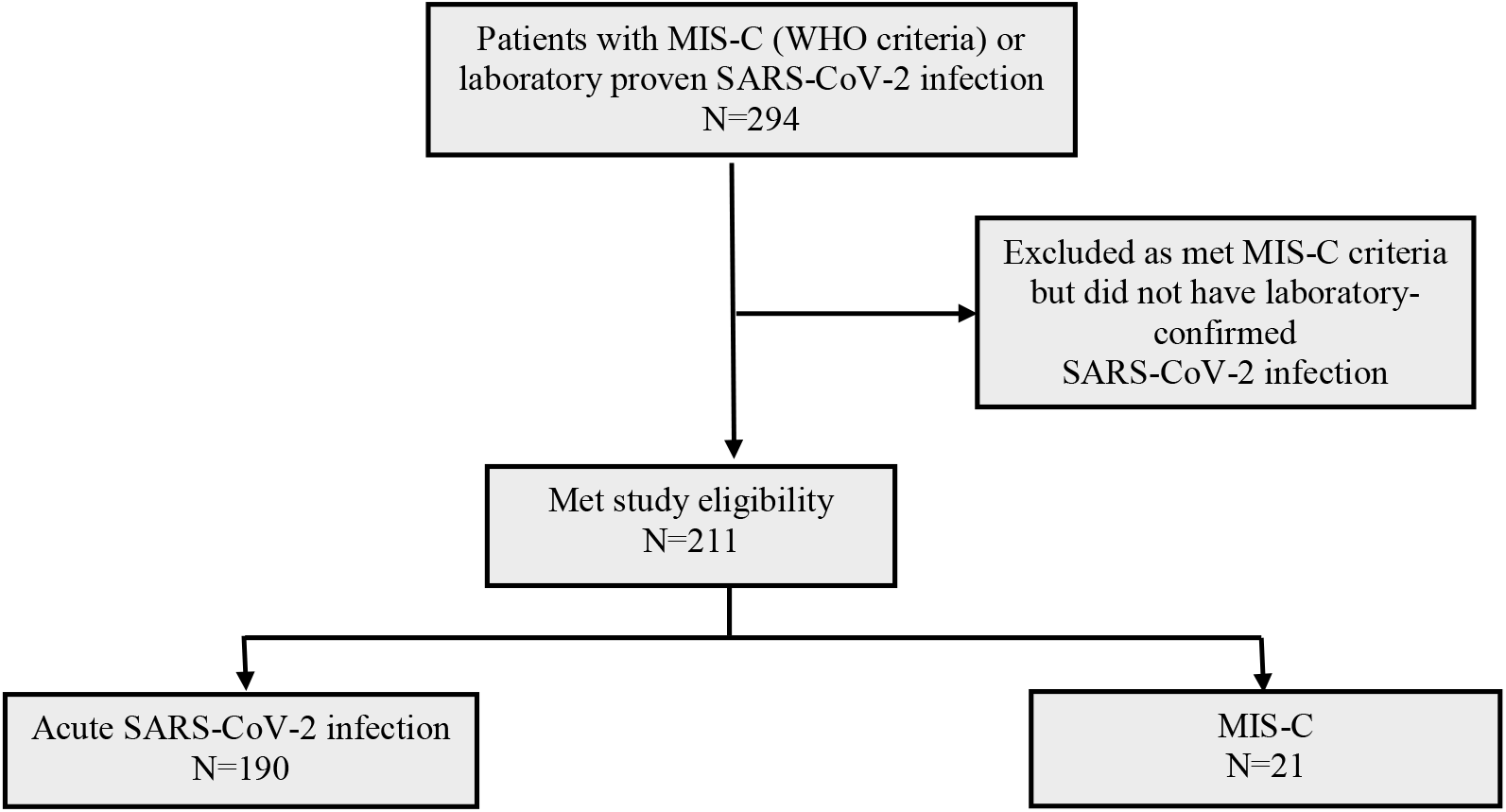
Flow diagram of patient enrollment.

### Comorbidities

Eighty-seven (41%) of the 211 children had at least one comorbidity (Table 1), including asthma (N=15; 7%), obesity (N=13; 6%), cancer (N=12; 6%), and other conditions requiring immunosuppressive drugs (N=10; 5%). Thirty-five (17%) had multiple comorbidities. Fourteen (7%) met the definition for medical complexity.

### Laboratory confirmation

Laboratory confirmation for the 211 patients was via molecular detection (N=185), serological confirmation (n=21) or both (N=5). Molecular detection was from the nasopharynx (N=154), nose (N=18), endotracheal tube (N=4), and throat (N=20) with some having detection from more than one site.

### Clinical features

Ages and symptoms are shown in Figures 2 and 3 respectively for patients with (N=21) and without (N=190) MIS-C. Fever was the most common symptom in both groups. Children with MIS-C were more likely to have fever, gastrointestinal symptoms, myalgias, conjunctivitis, cracked lips and rash (P<0.05) than were those without MIS-C. The median duration of fever was 7 days (interquartile range [IQR] 5, 9) and 3 days (IQR: 1, 5; P<0.001) in those with and without MIS-C respectively.

**Figure 2.**
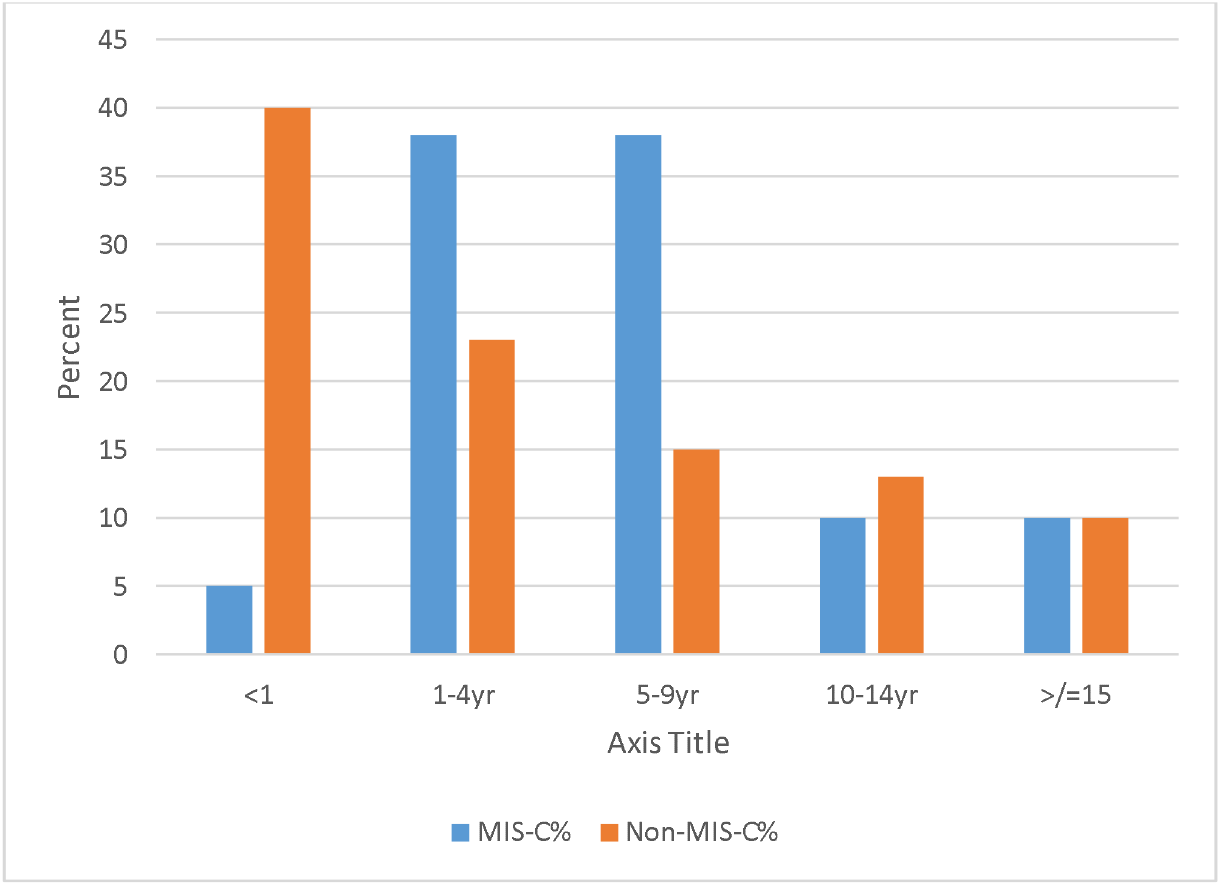
Age distribution of hospitalized children with laboratory-confirmed SARS-CoV-2 infection

**Figure 3.**
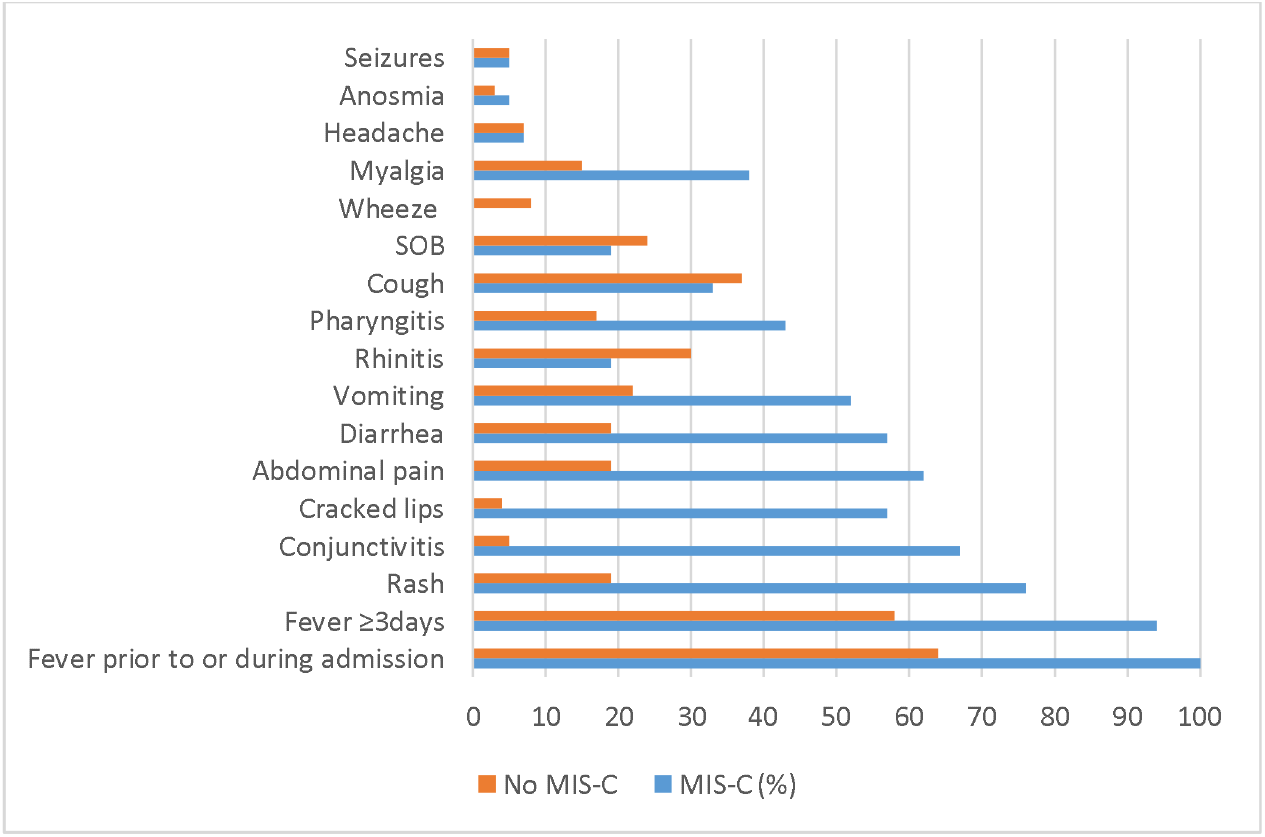
Frequency of signs and symptoms prior to or during hospitalization for 211 children with proven SARS-CoV-2 infection

### Role of SARS-Cov-2 in admission

One hundred eight of the 211 cases (51%) were not suspected to have COVID-19 or MIS-C on admission (Table 1). Eighty-one (76%) had asymptomatic or mild disease. As mentioned, 8 later developed healthcare-associated SARS-CoV-2.

### Coinfections

There were 23 bacterial and 16 viral coinfections (Table 1) with a wide variety of pathogens. Antibiotics were administered to 147 (70%) of the 211 children −36 (17%) for possible bacterial pneumonia and 111 (53%) for other indications. Of the 103 children admitted for suspected COVID-19 or MIS-C, 73 (71%) received antibiotics, of which 26 (25%) received them for possible bacterial pneumonia and 47 (46%) for other indications, often fever without a focus.

### Outcomes

Of the 211 cases, 140 (66%) did not require supplemental oxygen or ICU admission, 30 (14%) required supplemental oxygen but not ICU admission, one (0.5%) developed healthcare-associated COVID-19 while already in ICU, and 7 (3%) tested positive for SARS-CoV-2 on the day that they were admitted from home to ICU but the ICU admission was for other indications. The remaining 33 (16%) required ICU admission for COVID-19 (N=26) or MIS-C (N=7; 33% of MIS-C cases), of which 18 (55%) required mechanical ventilation for COVID-19 (N=14) or MIS-C (N=4; 19% of MIS-C cases). Of the medically complex children, 7 of 14 (50%) were admitted to ICU. This included the only child with a tracheostomy and the only child mechanically ventilated at home, 2 of 5 who were tube fed, 2 of 3 who were on home oxygen, and 1 of 4 who were on non-invasive ventilation on home. There were four deaths (three in Iran and one in Costa Rica): i) a child with a single kidney and ureteric reflux died of respiratory failure 5 days after admission with COVID-19, ii) a child admitted for chemotherapy for leukemia had healthcare-associated COVID-19 confirmed on day 21 of admission and died on day 34. iii) a child admitted with relapsed leukemia had healthcare-associated COVID-19 confirmed on day 16 of admission and died on day 19, and iv) a child with an untreatable brain tumour died 2 days after admission with COVID-19. SARS-CoV-2 was thought to have caused the first two deaths and hastened the latter two.

### Comparison of children admitted in Canada to those admitted in other countries

Table 2 compares the 95 children admitted in Canada to 84 admitted in Costa Rica and 32 admitted in Iran. Combining the data for the children from the middle-income countries (Costa Rica and Iran), children admitted in Canada were older (median 4.4 versus 2.2 years; p<0.05). Fever occurred in 51 of 82 children in Canada (data missing for 13 cases) versus 92 of 116 at other sites (62% vs 79%); p =0.008). Abdominal pain in children over 1 year of age (since one cannot reliably determine if infants have abdominal pain) was also less common at the Canadian sites (15 of 70 (23%) versus 35 of 69 (51%); p<0.001). Children admitted to Canadian sites had less severe illness. Although ICU admission rates were comparable (12/95 [13%] versus 21/ 116 [18%]; p =0.27), 3 (25%) of the 12 ICU admissions in Canada required mechanical ventilation versus 15 of 21 at the other sites (71%; p =0.014). Median hospital stay was shorter at Canadian sites (3 days versus 5 days; p<0.001). MIS-C criteria were met by 5 cases in Canada (5%) and 16 cases at other sites (13 in Costa Rica and 3 in Iran) (14%; p= 0.06). The COVID-19 cases in Iran mainly occurred early in the pandemic so were commonly managed with therapies that are no longer recommended (azithromycin, hydroxychloroquine, and lopinavir-ritonavir).

**Table 2.**
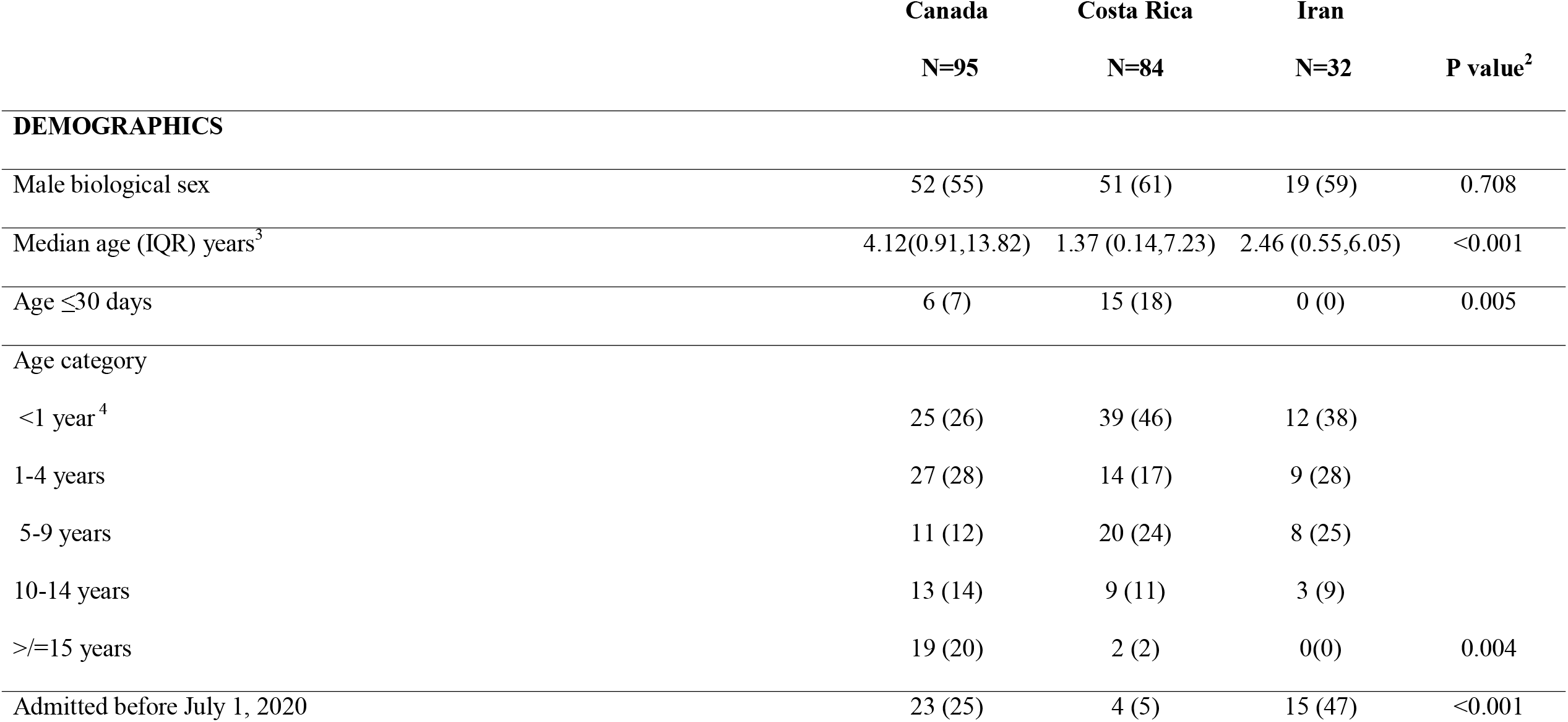
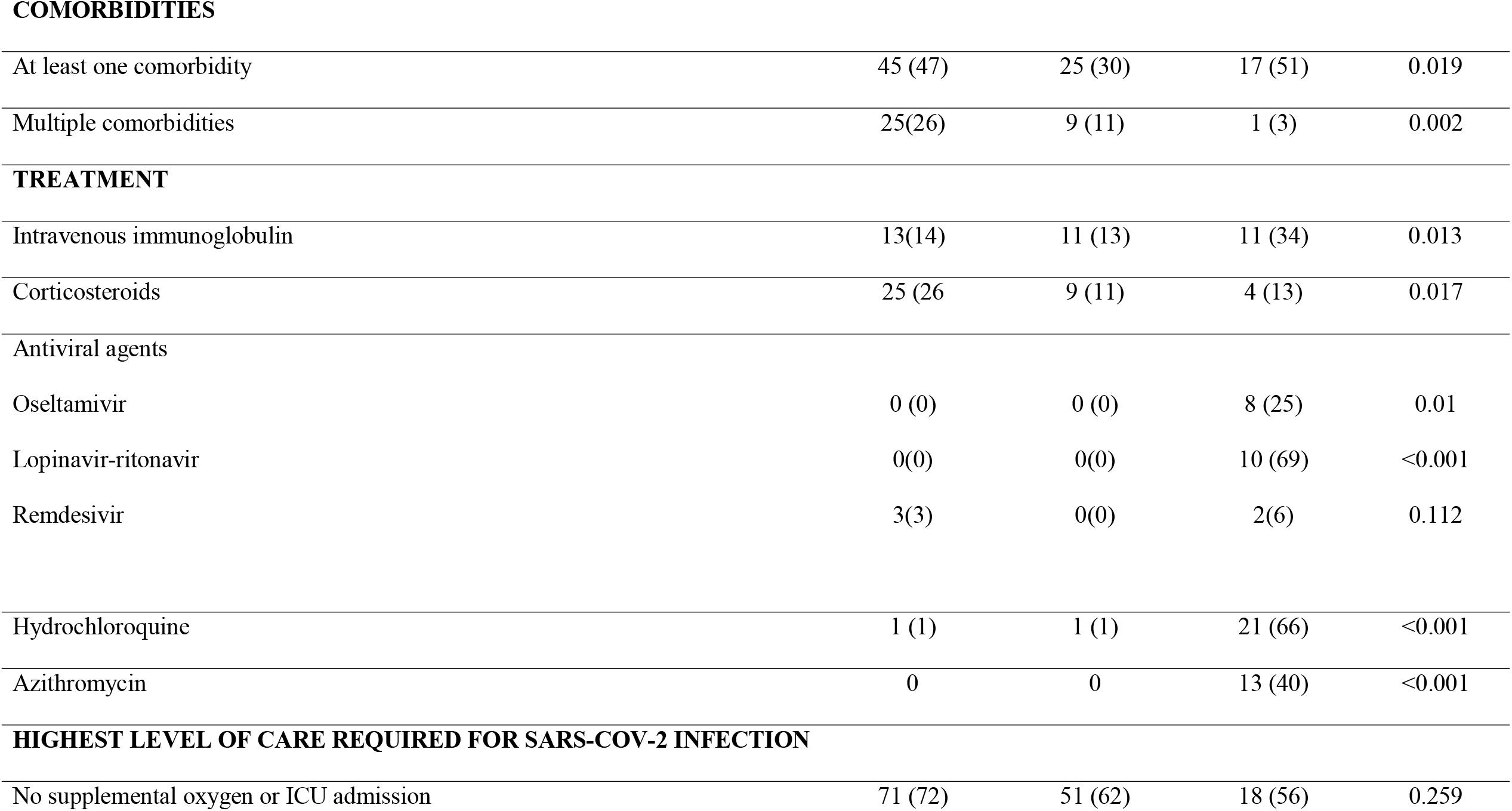
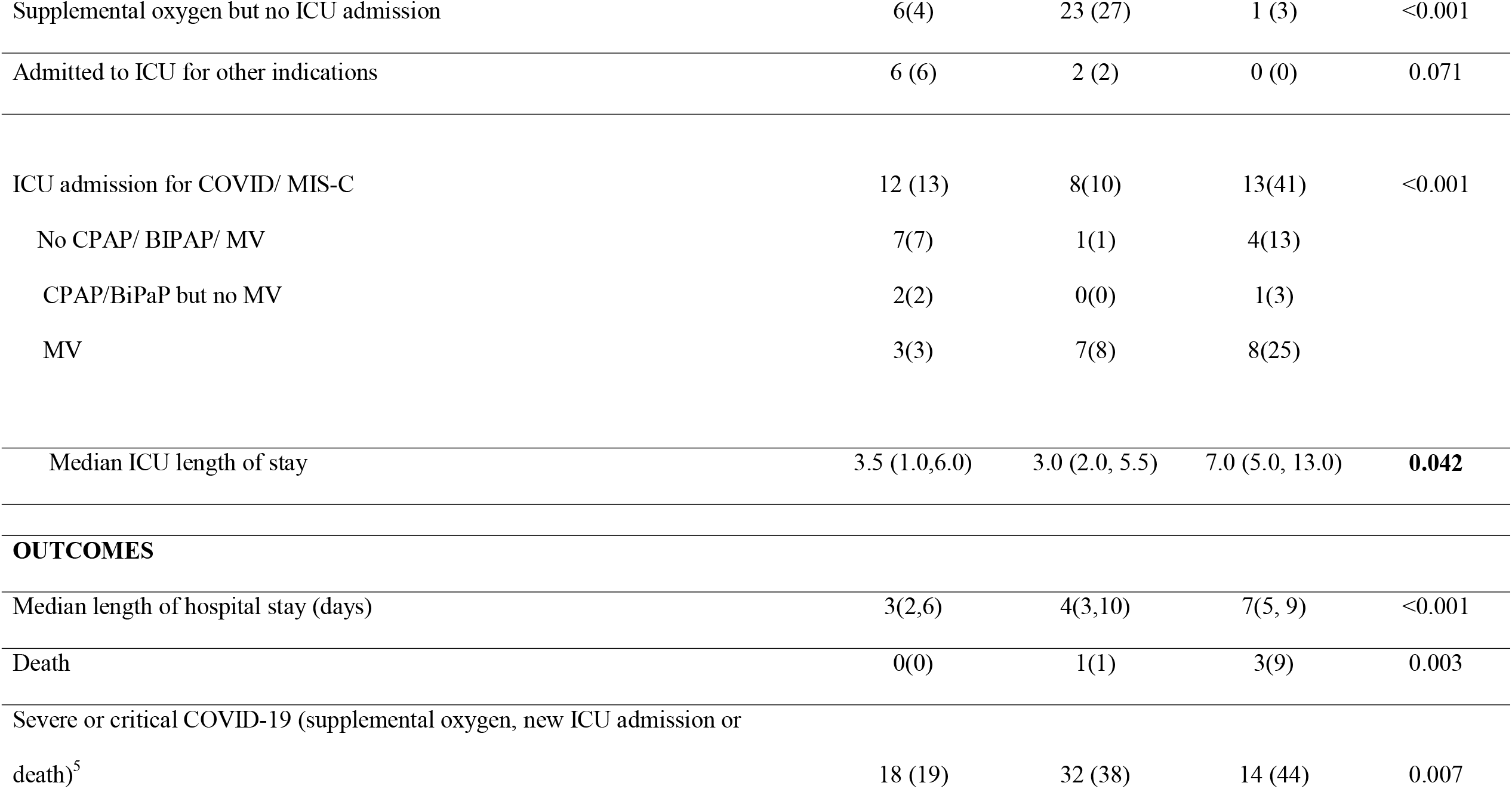
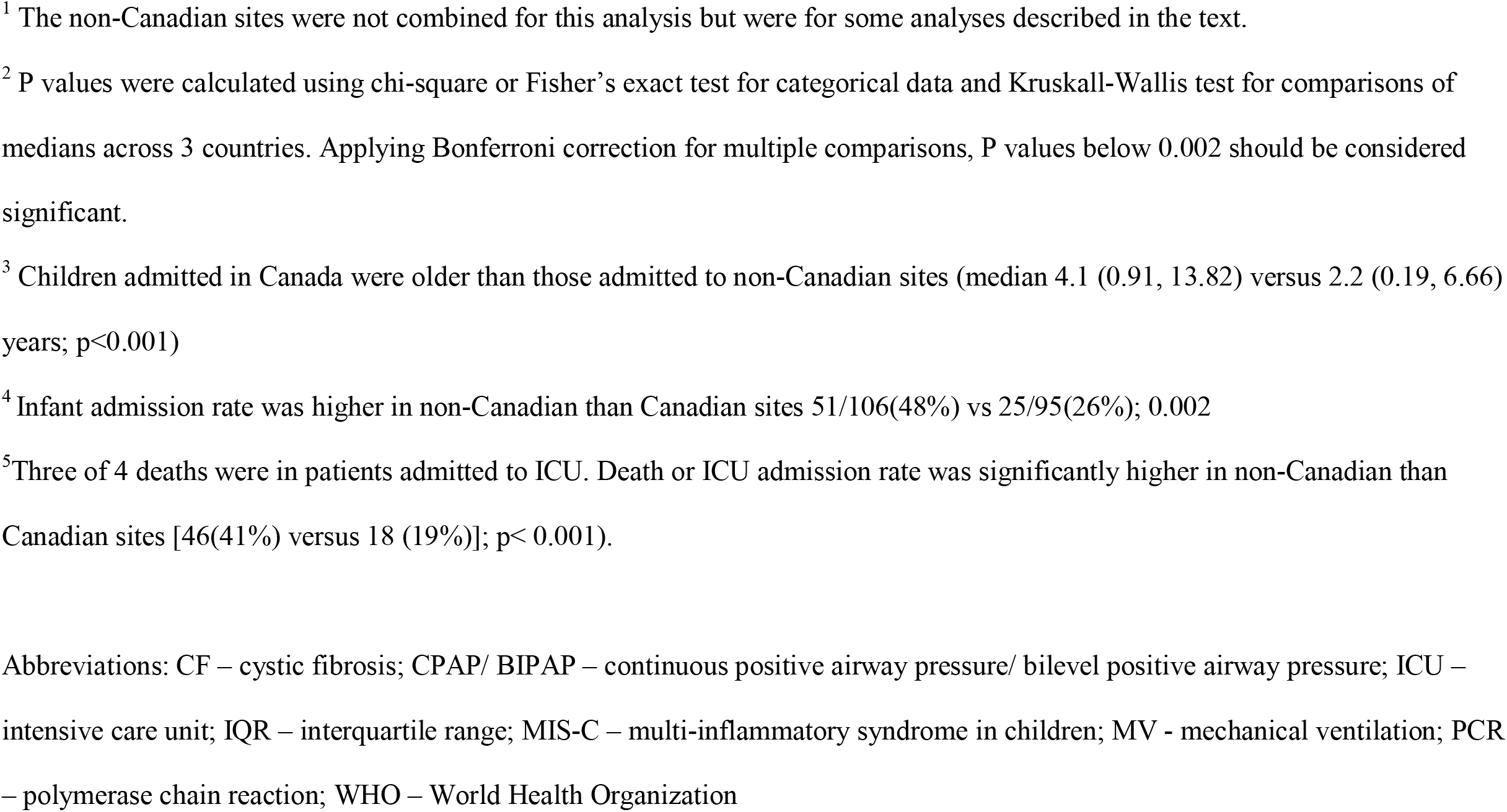
Comparison of 95 children with COVID-19 or MIS-C admitted in Canada with 116 children admitted in other countries^1^

|  | **Canada** | **Costa Rica** | **Iran** |  |
| --- | --- | --- | --- | --- |
|  | **N=95** | **N=84** | **N=32** | **P value<sup>2</sup>** |
| **DEMOGRAPHICS** |  |  |  |  |
| Male biological sex | 52 (55) | 51 (61) | 19 (59) | 0.708 |
| Median age (IQR) years<sup>3</sup> | 4.12(0.91,13.82) | 1.37 (0.14,7.23) | 2.46 (0.55,6.05) | <0.001 |
| Age ≤30 days | 6 (7) | 15 (18) | 0 (0) | 0.005 |
| Age category |  |  |  |  |
| <1 year<sup>4</sup> | 25 (26) | 39 (46) | 12 (38) |  |
| 1-4 years | 27 (28) | 14 (17) | 9 (28) |  |
| 5-9 years | 11 (12) | 20 (24) | 8 (25) |  |
| 10-14 years | 13 (14) | 9 (11) | 3 (9) |  |
| ≥15 years | 19 (20) | 2 (2) | 0(0) | 0.004 |
| Admitted before July 1, 2020 | 23 (25) | 4 (5) | 15 (47) | <0.001 |
| **COMORBIDITIES** |  |  |  |  |
| At least one comorbidity | 45 (47) | 25 (30) | 17 (51) | 0.019 |
| Multiple comorbidities | 25(26) | 9 (11) | 1 (3) | 0.002 |
| **TREATMENT** |  |  |  |  |
| Intravenous immunoglobulin | 13(14) | 11 (13) | 11 (34) | 0.013 |
| Corticosteroids | 25 (26 | 9 (11) | 4 (13) | 0.017 |
| Antiviral agents |  |  |  |  |
| Oseltamivir | 0 (0) | 0 (0) | 8 (25) | 0.01 |
| Lopinavir-ritonavir | 0(0) | 0(0) | 10 (69) | <0.001 |
| Remdesivir | 3(3) | 0(0) | 2(6) | 0.112 |
| Hydrochloroquine | 1 (1) | 1 (1) | 21 (66) | <0.001 |
| Azithromycin | 0 | 0 | 13 (40) | <0.001 |
| **HIGHEST LEVEL OF CARE REQUIRED FOR SARS-COV-2 INFECTION** |  |  |  |  |
| No supplemental oxygen or ICU admission | 71 (72) | 51 (62) | 18 (56) | 0.259 |
| Supplemental oxygen but no ICU admission | 6(4) | 23 (27) | 1 (3) | <0.001 |
| Admitted to ICU for other indications | 6 (6) | 2 (2) | 0 (0) | 0.071 |
| ICU admission for COVID/ MIS-C | 12 (13) | 8(10) | 13(41) | <0.001 |
| No CPAP/ BIPAP/ MV | 7(7) | 1(1) | 4(13) |  |
| CPAP/BiPaP but no MV | 2(2) | 0(0) | 1(3) |  |
| MV | 3(3) | 7(8) | 8(25) |  |
| Median ICU length of stay | 3.5 (1.0,6.0) | 3.0 (2.0, 5.5) | 7.0 (5.0, 13.0) | **0.042** |
| **OUTCOMES** |  |  |  |  |
| Median length of hospital stay (days) | 3(2,6) | 4(3,10) | 7(5, 9) | <0.001 |
| Death | 0(0) | 1(1) | 3(9) | 0.003 |
| Severe or critical COVID-19 (supplemental oxygen, new ICU admission or death)<sup>5</sup> | 18 (19) | 32 (38) | 14 (44) | 0.007 |

<sup>1</sup> The non-Canadian sites were not combined for this analysis but were for some analyses described in the text.
<sup>2</sup> P values were calculated using chi-square or Fisher's exact test for categorical data and Kruskal-Wallis test for comparisons of medians across 3 countries. Applying Bonferroni correction for multiple comparisons, P values below 0.002 should be considered significant.
<sup>3</sup> Children admitted in Canada were older than those admitted to non-Canadian sites (median 4.1 (0.91, 13.82) versus 2.2 (0.19, 6.66) years; $p < 0.001$ )
<sup>4</sup> Infant admission rate was higher in non-Canadian than Canadian sites 51/106(48%) vs 25/95(26%); 0.002
<sup>5</sup> Three of 4 deaths were in patients admitted to ICU. Death or ICU admission rate was significantly higher in non-Canadian than Canadian sites [46(41%) versus 18 (19%)]; $p < 0.001$ ).
Abbreviations: CF – cystic fibrosis; CPAP/ BIPAP – continuous positive airway pressure/ bilevel positive airway pressure; ICU – intensive care unit; IQR – interquartile range; MIS-C – multi-inflammatory syndrome in children; MV - mechanical ventilation; PCR – polymerase chain reaction; WHO – World Health Organization

### Risk factors for severe or critical COVID-19

In the multivariate analysis, age ≤30 days, hospitalization outside of Canada, the presence of at least one comorbidity and chest imaging abnormalities compatible with COVID-19 remained significant (Table 3).

**Table 3:**
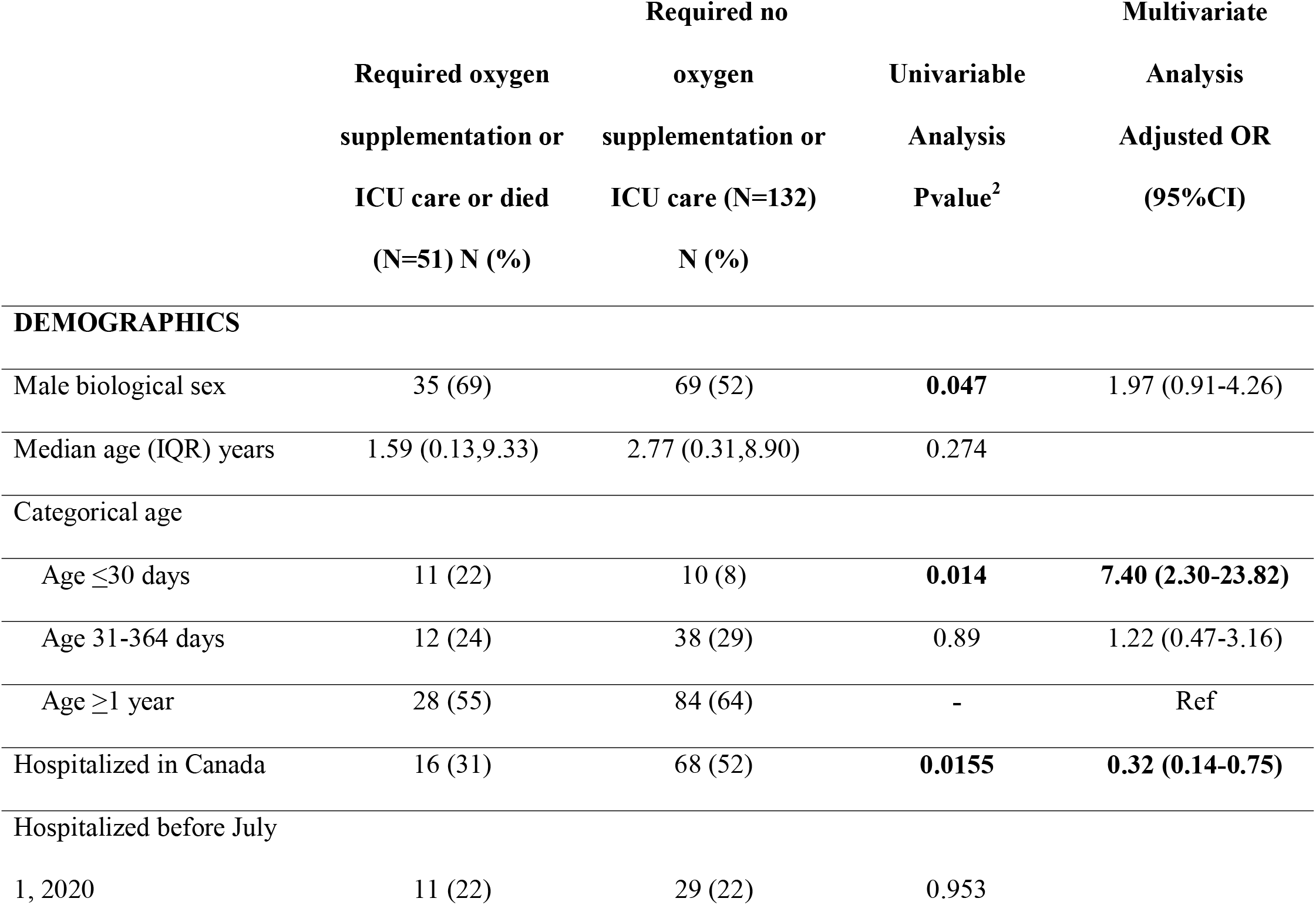
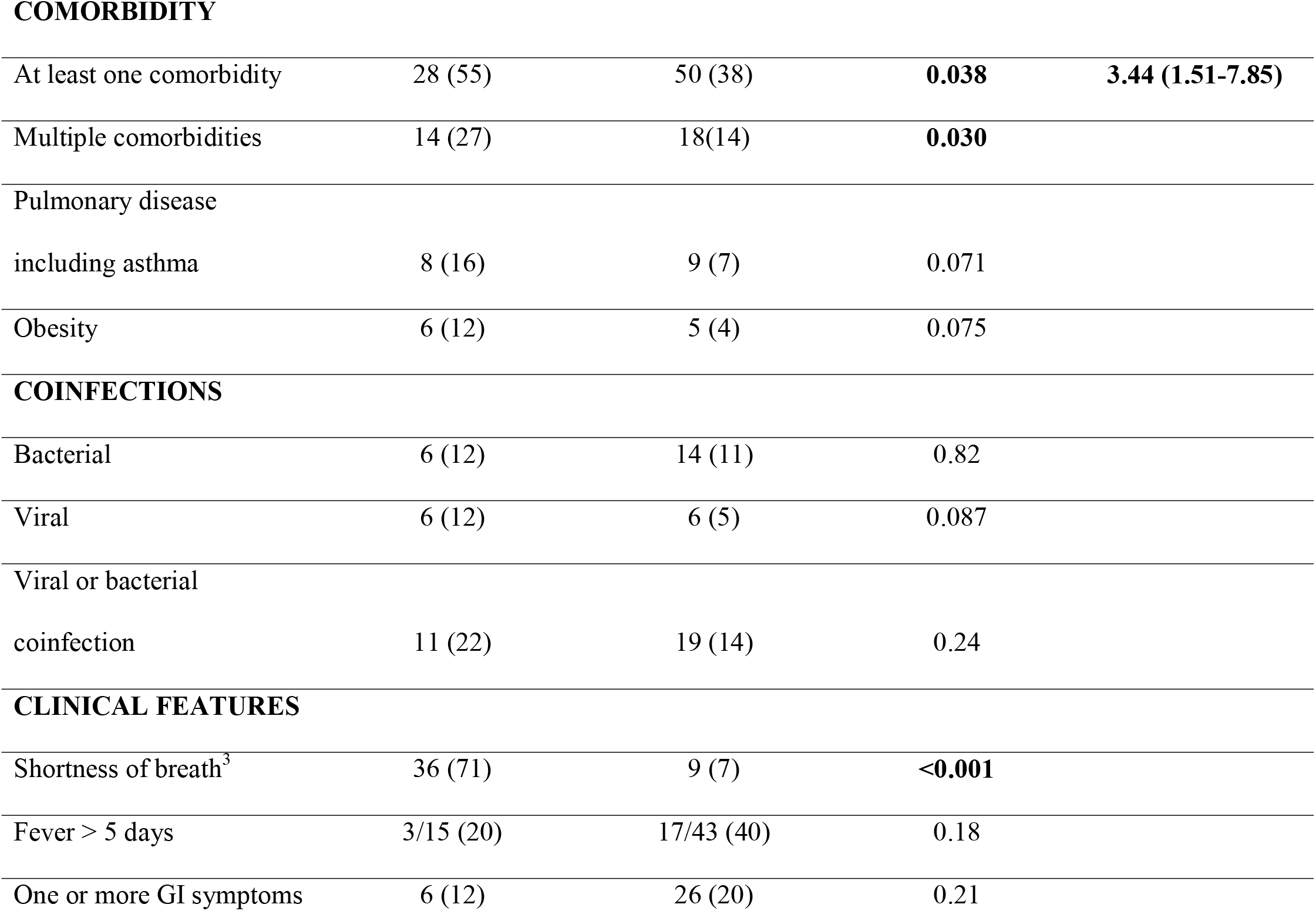
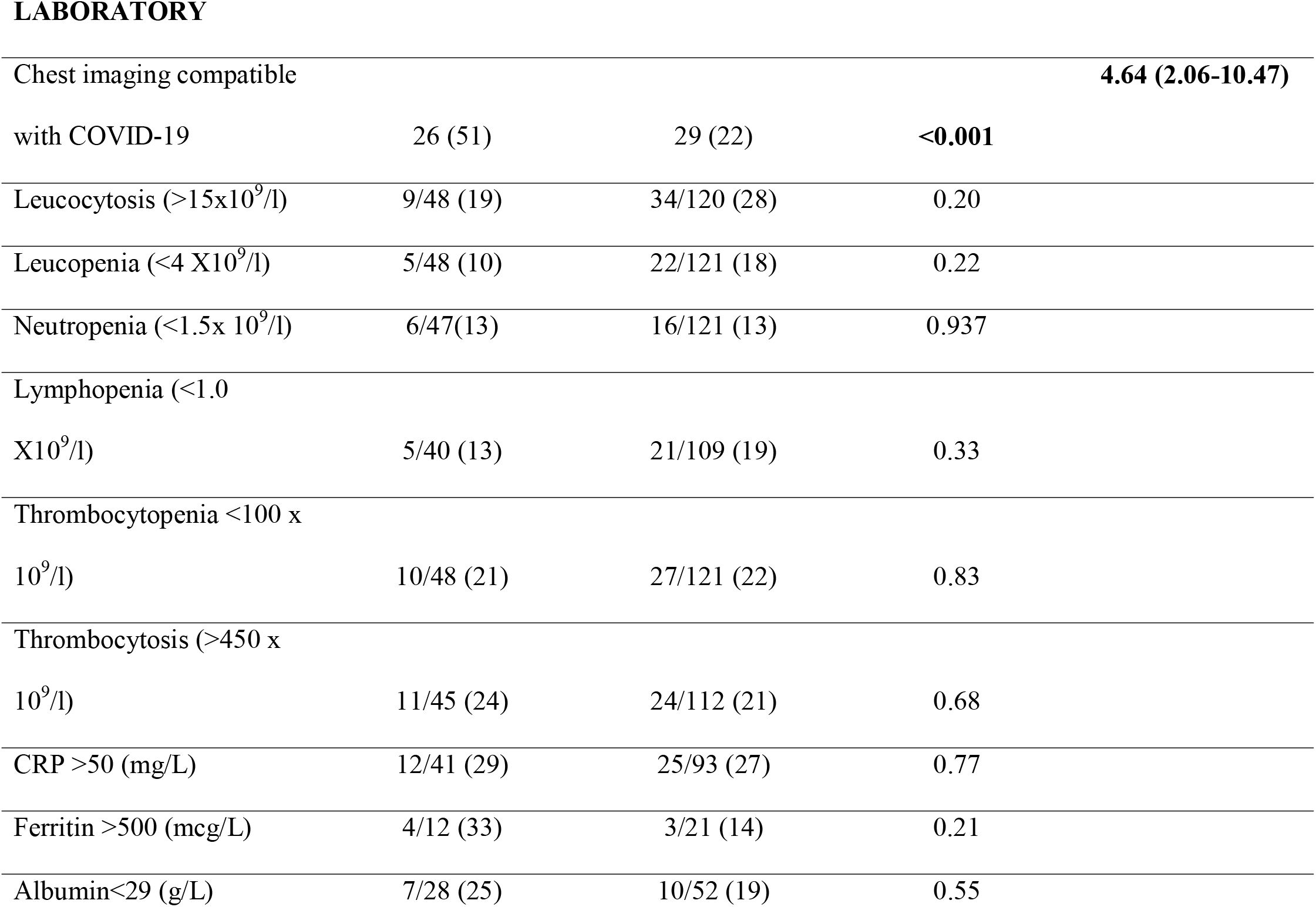
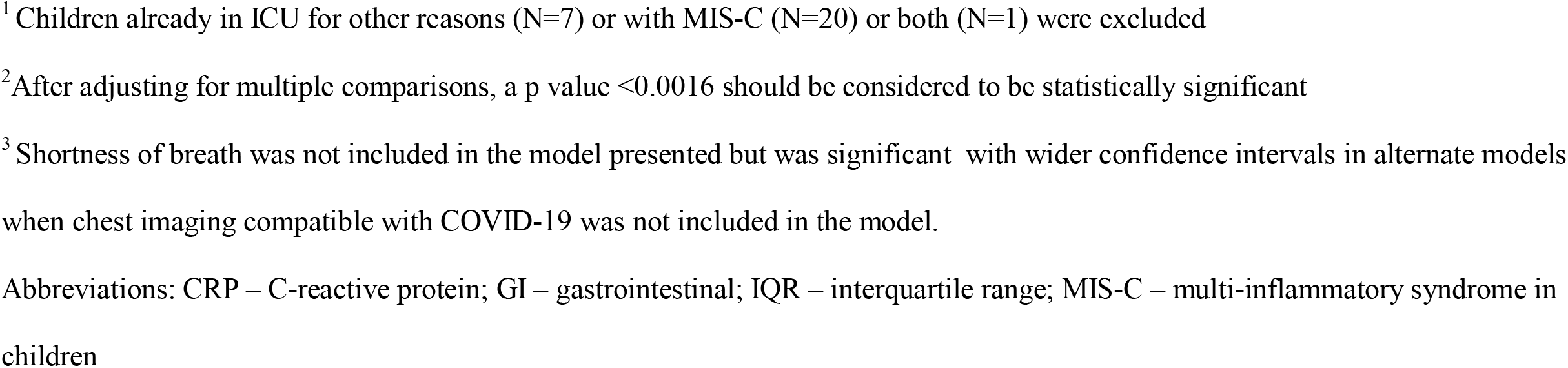
Risk Factors for severe or critical disease in 183 children hospitalized with COVID-19^1^

|  | Required oxygen<br>supplementation or<br>ICU care or died<br>(N=51) N (%) | Required no<br>oxygen<br>supplementation or<br>ICU care (N=132)<br>N (%) | Univariable<br>Analysis<br>Pvalue<sup>2</sup> | Multivariate<br>Analysis<br>Adjusted OR<br>(95%CI) |
| --- | --- | --- | --- | --- |
| **DEMOGRAPHICS** |  |  |  |  |
| Male biological sex | 35 (69) | 69 (52) | **0.047** | 1.97 (0.91-4.26) |
| Median age (IQR) years | 1.59 (0.13,9.33) | 2.77 (0.31,8.90) | 0.274 |  |
| Categorical age |  |  |  |  |
| Age ≤30 days | 11 (22) | 10 (8) | **0.014** | **7.40 (2.30-23.82)** |
| Age 31-364 days | 12 (24) | 38 (29) | 0.89 | 1.22 (0.47-3.16) |
| Age ≥1 year | 28 (55) | 84 (64) | - | Ref |
| Hospitalized in Canada | 16 (31) | 68 (52) | **0.0155** | **0.32 (0.14-0.75)** |
| Hospitalized before July |  |  |  |  |
| 1, 2020 | 11 (22) | 29 (22) | 0.953 |  |
| **COMORBIDITY** |  |  |  |  |
| At least one comorbidity | 28 (55) | 50 (38) | **0.038** | **3.44 (1.51-7.85)** |
| Multiple comorbidities | 14 (27) | 18(14) | **0.030** |  |
| Pulmonary disease |  |  |  |  |
| including asthma | 8 (16) | 9 (7) | 0.071 |  |
| Obesity | 6 (12) | 5 (4) | 0.075 |  |

**COINFECTIONS**
|  |  |  |  |
| --- | --- | --- | --- |
| Bacterial | 6 (12) | 14 (11) | 0.82 |
| Viral | 6 (12) | 6 (5) | 0.087 |
| Viral or bacterial |  |  |  |
| coinfection | 11 (22) | 19 (14) | 0.24 |

**CLINICAL FEATURES**
|  |  |  |  |
| --- | --- | --- | --- |
| Shortness of breath<sup>3</sup> | 36 (71) | 9 (7) | **<0.001** |
| Fever > 5 days | 3/15 (20) | 17/43 (40) | 0.18 |
| One or more GI symptoms | 6 (12) | 26 (20) | 0.21 |

LABORATORY
|  |  |  |  |
| --- | --- | --- | --- |
| Chest imaging compatible |  |  | **4.64 (2.06-10.47)** |
| with COVID-19 | 26 (51) | 29 (22) | **<0.001** |
| Leucocytosis ($>15 \times 10^9/l$) | 9/48 (19) | 34/120 (28) | 0.20 |
| Leucopenia ($<4 \times 10^9/l$) | 5/48 (10) | 22/121 (18) | 0.22 |
| Neutropenia ($<1.5 \times 10^9/l$) | 6/47(13) | 16/121 (13) | 0.937 |
| Lymphopenia ($<1.0 \times 10^9/l$) | 5/40 (13) | 21/109 (19) | 0.33 |
| Thrombocytopenia $<100 \times 10^9/l$) | 10/48 (21) | 27/121 (22) | 0.83 |
| Thrombocytosis ($>450 \times 10^9/l$) | 11/45 (24) | 24/112 (21) | 0.68 |
| CRP $>50$ (mg/L) | 12/41 (29) | 25/93 (27) | 0.77 |
| Ferritin $>500$ (mcg/L) | 4/12 (33) | 3/21 (14) | 0.21 |
| Albumin$<29$ (g/L) | 7/28 (25) | 10/52 (19) | 0.55 |

<sup>1</sup> Children already in ICU for other reasons (N=7) or with MIS-C (N=20) or both (N=1) were excluded
<sup>2</sup> After adjusting for multiple comparisons, a p value <0.0016 should be considered to be statistically significant
<sup>3</sup> Shortness of breath was not included in the model presented but was significant with wider confidence intervals in alternate models when chest imaging compatible with COVID-19 was not included in the model.
Abbreviations: CRP – C-reactive protein; GI – gastrointestinal; IQR – interquartile range; MIS-C – multi-inflammatory syndrome in children

## Discussion

Of 211 children hospitalized with documented SARS-CoV-2 infection with or without MIS-C, approximately half were admitted with other suspected diagnoses and approximately two-thirds had asymptomatic or mild disease. However, 33 (16%) required ICU admission due to COVID-19 or MIS-C of whom about half required mechanical ventilation. Four died (1.9%) with three of the deaths occurring in children with malignancy.

Similar to other cohort studies conducted in developed countries, the minority of patients in this current study (30%) required supplemental oxygen therapy or ICU admission. Of 363 children admitted to 82 health care institutions in Europe in April 2020, 75 (21%) required supplemental oxygen, including 48 (13%) who required ICU admission; 25 (7%) were managed with mechanical ventilation and 4 (1.1%) died [15]. Of 281 patients up to 22 years of age admitted to 8 US hospitals up to April 12, 2020, 70 of 212 (33%) with COVID-19 were admitted to ICU, 26 (12%) required mechanical ventilation and 7 (3%) died while 44 of 69 (64%) with MIS-C were admitted to ICU, 3 (4%) required mechanical ventilation and none died [16].

The incidence and burden of asymptomatic and mild infections in hospitalized children is just beginning to be described; 34 of 315 infected youth and young adults (11%) admitted to 8 United States (US) hospitals in April 2020 had “incidental” SARS-CoV-2 infection [16]. SARS-CoV-2 was not suspected at the time of admission in approximately half of the children in the current study. Some centers now routinely screen all children at admission for SARS-CoV-2. However, approximately 5% of cases in the current study were acquired in the hospital so would not have been detected by this strategy. Our methodology prevented us from determining whether children with SARS-CoV-2 infection were appropriately placed on additional precautions. The possibility of SARS-CoV-2 infection needs to be considered in all admitted children until the pandemic ends. Although it remains uncertain how often infected children transmit to others, the need to isolate children with possible and proven SARS-CoV-2 infection poses a significant and increasing burden in pediatric hospitals. Moreover, caregivers ought to also be considered as potential vectors of SARS-CoV-2 transmission in the pediatric hospital setting.

It is now well recognized that comorbidities increase the risk of hospitalization for children with SARS-CoV-2 infection. A US series reported that 94 of 244 (42%) admitted children had comorbidities, with obesity being the most common ([17]. In the current study, approximately 40% of admitted children had comorbidities but our methodology did not allow us to determine the risk of hospitalization for any given comorbidity. Establishment of risk factors for hospitalization is of potential relevance as the highest risk patients may eventually be suitable targets for immunization or novel early therapies [18].

Identifying risk factors for severe disease in hospitalized children is perhaps of even greater importance as novel early therapies become available. In the current study, neonatal age group, hospitalization outside of Canada, the presence of at least one comorbidity and chest imaging abnormalities compatible with COVID-19 were risk factors for severe or critical COVID-19. A systematic review of 285 014 children with SARS-CoV-2 infection reported that 5.1% with comorbidities and 0.2% without developed severe disease [5] but as with hospitalization, the risk with any given comorbidity is unknown. A study of 48 patients up to age 21 years admitted to ICUs in the United States reported that 40 (83%) had comorbidities (versus 56% in the current study) of which 19 were medically complex (defined as long-term dependence on technological support associated with developmental delay and/or genetic abnormalities) [6]. In the current study, 7 of the 33 children (21%) admitted to ICU due to COVID-19 fulfilled a similar definition of medical complexity. In the previously mentioned European study, risk factors for ICU admission with either COVID-19 or MIS-C included age younger than 1-month, male sex, comorbidities, and presence of lower respiratory tract infection signs or symptoms at presentation (not further defined) [15]. In the previously mentioned study from the US, risk factors for ICU admission for 48 hours of more for COVID-19 included younger age, obesity, hypoxia, higher admission white blood cell count and bilateral infiltrates on admission chest radiograph [16]; risk factors for ICU admission for MIS-C were not reported. A previous pediatric systematic review also reported obesity as a risk factor for severe disease [19] while it did not appear to be a risk factor in the current study. In a study from Oman, leukocytosis for age, elevated CRP (optimal cut-off was >100 mg/L), and anemia for age were risk factors for ICU admission for COVID-19 or MIS-C [20].

Cases of SARS-Co-2 infection in non-Canadian sites appeared to have a greater severity of illness with a higher prevalence of MIS-C cases. It is possible that the threshold for admission or for testing is lower in Canada or that children present for medical attention later in the course of illness in other countries. There may also be a genetic component as certain ethnic groups appear to be at greater risk for MIS-C [21]. Thus, the incidence may vary considerably by country.

This study has the inherent limitations of a retrospective chart review. It was not population based as most participating hospitals are referral centers so milder cases might have been admitted elsewhere. Indications for testing for SARS-CoV-2 and available assays varied by center. We were not able to account for the severity of comorbidities. Interpretation of chest radiographs was not standardized. Prior studies suggest that Black and Latin American children are at increased risk of COVID-19 [22] and of MIS-C [21] but race or ethnic background were not available in our cohort. MIS-C cases are under-reported as i) MIS-C was yet to be described when the study commenced so relevant clinical questions were not included in the original version of the case report form and ii) SARS-CoV-2 serology was not available at any sites early in the study and remained unavailable by the end of the study at some sites.

In conclusion, approximately half of children with SARS-CoV-2 infection or MIS-C diagnosed during hospitalization were admitted with another diagnosis. Clinicians must consider the diagnosis of community-acquired or healthcare-associated SARS-CoV-2 infection in all admitted children. Neonates, children with comorbidities and those with a chest radiograph compatible with COVID-19 were at highest risk of requiring supplemental oxygen or ICU admission during SARS-CoV-2 hospitalization. Death was rare, but children with malignancies appear to be over-represented. Future studies are needed to examine socio-demographic factors that may drive hospitalization rates and outcomes related to SARS-CoV-2 infection in children.

## Data Availability

Data are available from the corresponding author.

## References

1. Hopkins J. Coronavirus Resource Center Global Map 2020.

2. Dong Y, Mo X, Hu Y, et al. Epidemiology of COVID-19 Among Children in China. Pediatrics 2020; 145(6).

3. Tezer H, Bedir Demirdağ T. Novel coronavirus disease (COVID-19) in children. Turkish journal of medical sciences 2020; 50(Si-1): 592–603.

4. Coronavirus Disease 2019 in Children - United States, February 12-April 2, 2020. MMWR Morbidity and mortality weekly report 2020; 69(14): 422–6.

5. Tsankov BK, Allaire JM, Irvine MA, et al. Severe COVID-19 Infection and Pediatric Comorbidities: A Systematic Review and Meta-Analysis. International journal of infectious diseases : IJID : official publication of the International Society for Infectious Diseases 2020.

6. Shekerdemian LS, Mahmood NR, Wolfe KK, et al. Characteristics and Outcomes of Children With Coronavirus Disease 2019 (COVID-19) Infection Admitted to US and Canadian Pediatric Intensive Care Units. JAMA pediatrics 2020; 174(9): 868–73.

7. Oran DP, Topol EJ. The Proportion of SARS-CoV-2 Infections That Are Asymptomatic : A Systematic Review. Annals of internal medicine 2021.

8. Alsharrah D, Alhaddad F, Alyaseen M, et al. Clinical characteristics of pediatric SARS-CoV-2 infection and coronavirus disease 2019 (COVID-19) in Kuwait. Journal of medical virology 2020.

9. Organization WH. Multisystem inflammatory syndrome in children and adolescents temporally related to COVID-19. 2020.

10. Kaushik A, Gupta S, Sood M, Sharma S, Verma S. A Systematic Review of Multisystem Inflammatory Syndrome in Children Associated With SARS-CoV-2 Infection. The Pediatric infectious disease journal 2020; 39(11): e340–e6.

11. Bank TW. World Bank Country and Lending Groups.

12. Cuschieri S. The STROBE guidelines. Saudi journal of anaesthesia 2019; 13(Suppl 1): S31–s4.

13. Program CNIS. Surveillance for COVID-19 and other viral respiratory infections among inpatients in CNISP hospitals 2020; (Version: December 7, 2020).

14. Horwitz LI, Jones SA, Cerfolio RJ, et al. Trends in COVID-19 Risk-Adjusted Mortality Rates. Journal of hospital medicine 2020.

15. Götzinger F, Santiago-García B, Noguera-Julián A, et al. COVID-19 in children and adolescents in Europe: a multinational, multicentre cohort study. The Lancet Child & adolescent health 2020; 4(9): 653–61.

16. Fernandes DM, Oliveira CR, Guerguis S, et al. Severe Acute Respiratory Syndrome Coronavirus 2 Clinical Syndromes and Predictors of Disease Severity in Hospitalized Children and Youth. The Journal of pediatrics 2020.

17. Kim L, Whitaker M, O’Halloran A, et al. Hospitalization Rates and Characteristics of Children Aged <18 Years Hospitalized with Laboratory-Confirmed COVID-19 - COVID-NET, 14 States, March 1-July 25, 2020. MMWR Morbidity and mortality weekly report 2020; 69(32): 1081–8.

18. Wolf J, Abzug MJ, Wattier RL, et al. Initial Guidance on Use of Monoclonal Antibody Therapy for Treatment of COVID-19 in Children and Adolescents. Journal of the Pediatric Infectious Diseases Society 2021.

19. Hoang A, Chorath K, Moreira A, et al. COVID-19 in 7780 pediatric patients: A systematic review. EClinicalMedicine 2020; 24: 100433.

20. Al Yazidi LS, Al Hinai Z, Al Waili B, et al. Epidemiology, characteristics, and outcomes of hospitalized children with COVID-19 in Oman: A multicenter cohort study. International journal of infectious diseases : IJID : official publication of the International Society for Infectious Diseases 2021.

21. Rowley AH, Shulman ST, Arditi M. Immune pathogenesis of COVID-19-related multisystem inflammatory syndrome in children. The Journal of clinical investigation 2020; 130(11): 5619–21.

22. Leeb RT, Price S, Sliwa S, et al. COVID-19 Trends Among School-Aged Children - United States, March 1-September 19, 2020. MMWR Morbidity and mortality weekly report 2020; 69(39): 1410–5.

